# Effect of exercise and dietary intervention on serum metabolomics in men with insomnia symptoms: a 6-month randomized-controlled trial

**DOI:** 10.1101/2020.02.23.20026898

**Authors:** Xiaobo Zhang, Xiuqiang Wang, Shenglong Le, Xiaowei Ojanen, Xiao Tan, Petri Wiklund, Sulin Cheng

## Abstract

**Background:** Accumulating evidences have shown that lifestyle interventions such as exercise and diet are associated with improved sleep quality. However, the underlying molecular mechanisms remain unclear. Assessing exercise and diet intervention associated changes in circulating metabolomics profile in people with insomnia symptoms may help to identify molecular biomarkers that may link lifestyle changes to improved sleep outcomes.

**Methods:** The present study is a part of a 6-month randomized lifestyle intervention on sleep disorder subjects. Seventy-two Finnish men (aged: 51.6 ± 10.1 years; body mass index, BMI: 29.3 ± 3.9 kg/m^2^) with chronic insomnia symptoms who were assigned into different intervention groups completed this study (exercise n = 24, diet n = 27 and control n = 21). The exercise group was assigned to a progressive aerobic exercise training with intensity of 60 – 75% of estimated maximum heart rate, 3 – 5 times a week. The diet group aimed to reduce their total energy intakes by 300 to 500 kcal per day for the first three months. The control group were advised to maintain their current lifestyle. Sleep was assessed by using a non-contact sleep monitoring devise (Beddit sleep tracker). Blood samples were collected in the morning between 7:00 and 9:00 a.m. after overnight fasting. Gas Chromatography Time-Of-Flight Mass Spectrometry (GC-TOF-MS) method was used to determine the serum metabolites.

**Results:** Twenty-one metabolites were significantly changed in the exercise group, thirty-three metabolites in the diet group and five metabolites in the control group after intervention, respectively. The differential metabolites after exercise intervention were mainly related to glycerolipids and carbohydrates metabolism, while dietary intervention altered mainly amino acids metabolism and fatty acids metabolism related metabolites. We subsequently assessed the change of those metabolites with the change of sleep parameters and found that decreased alpha-ketoisocaproic acid (r = -0.52, p = 0.026) was correlated with improved sleep efficiency (SE) in the exercise group. Change of 3-hydroxybutric acid (r = -0.47, p = 0.025) and D-glucopyranose (r = -0.54, p = 0.006) correlated negatively with SE in the diet group. On the other hand, oxalic acid (r = 0.49, p = 0.021), D-glucopyranose (r = 0.43, p = 0.048), 4-deoxyerythronic acid (r = 0.60, p = 0.004) and tagatose (r = 0.51, p = 0.016) correlated positively with change of SOL, and 2-keto-isovaleric acid (r = 0.45, p = 0.029) correlated with TST in the diet group.

**Conclusion:** In conclusion, this study identified circulating metabolites that may represent a part of a biological mechanism through which lifestyle interventions are associated with improved sleep quality in people with insomnia.

## 1. Introduction

Insomnia, which is the most prevalent sleep disorder, is common in the modern society and have become an important public health issue ^1^. The prevalence of insomnia symptoms was 21.4% in USA adults assessed by DSM-IV criteria ^2^. Furthermore, sleep loss or sleep disruptions induced by insomnia symptoms are also associated with the increase in all-cause mortality and lead to detrimental effects on neuroendocrine systems ^1, 3^.

We have shown recently that six months exercise or diet intervention shortened objective sleep onset latency (SOL) and improved subjective sleep quality in overweight and obese men with insomnia symptoms, independent of weight loss ^4, 5^. Several human studies among insomniacs or individuals with other sleep disorders showed that exercise (including chronic and acute exercise) intervention have beneficial effects for sleep by increasing total sleep time (TST) and reducing wake after sleep onset (WASO) ^6-11^. The acute aerobic exercise improved sleep quality by increasing activity of ascending brain serotonergic, while dietary supplementation of tryptophan (the only serotonin precursor) can reduce sleep onset latency ^12^. The results of dietary intervention targeting patients with sleep disorders are conflicting ^13^, with two studies showing isocaloric high fat intake associated with significantly better sleep than other diets in human ^4, 14^. A large cross-sectional study from China showed an association between decreased sleep duration and increased fat intake in 2828 adults ^15^. However, the mechanisms underlying these associations between exercise/diet and sleep disorder, especially in insomnia remain largely are unclear.

Serum high-throughput metabolomics profiling is a feasible method to study the characteristic changes in small molecular metabolites in a pathophysiological state. Studies have shown that metabolites glucose, amino acid, fatty acid and tricarboxylic acid cycle intermediates levels differed in individuals with sleep disorders compared with healthy people ^16-18^ as well as in animal studies ^19, 20^. However, no study has examined the effect of lifestyle intervention on circulating metabolome in patients with insomnia symptoms.Thus, we conducted the present study to assess the associations between serum metabolites and sleep quality after a randomized controlled six-month exercise or diet intervention in men with insomnia symptoms.

## 2. Methods

### 2.1. Subjects

The study is a part of a 6-month randomized lifestyle intervention on middle-aged Finnish men with sleep disorders (including both insomnia and sleep apnea) ^21^. For this report, we only included 72 men aged 30–65 years who self-reported having suffered from chronic insomnia lasting three months or longer and completed the 6-month intervention ^21^. The subjects further filled a Vitalmed sleep questionnaire (which consists of all the questions in the Basic Nordic Sleep Questionnaire and the Epworth Sleepiness Scale (ESS)) were collected and reviewed by a physician. Of the qualified subjects and who completed the intervention, 24 subjects were allocated into exercise group, 27 subjects to diet group and 21 subjects to control group.

The study was approved by the Ethics Committee of the Central Finland Health Care District (7/2011 OTE) and registered under www.controlled-trials.com: ISRCTN77172005. Written informed consent was obtained from all participants before the baseline measurements and a copy of the signed consent form was archived.

### 2.2. Background information and anthropometric measurements

All lifestyle and medical history information were collected by self-administered questionnaires at the Laboratory of Sport and Health Sciences, University of Jyväskylä. Anthropometric measurements were performed after overnight fasting (12 h). Height was measured using a fixed wall scale device to the nearest 0.1 cm. Weight was determined to the nearest 0.1 kg using an electronic scale and calibrated before each measurement session. BMI was calculated as weight (kg) per height^2^ (m^2^).

### 2.3. Objective sleep measurement

Seven-night sleep measurements both before and after intervention were performed for all participants by using a non-contact sleep monitoring system at their homes (Beddit sleep tracker, Beddit.com Oy, Espoo, Finland) ^22^ and automatically analyzed by the Beddit server via internet. The system included a piezoelectric bed sensor. Ballistocardiographic signals were sampled by the piezoelectric sensor at 140 Hz and simultaneously uploaded to a web server through the Internet, where sleep/wake status was classified in 30-s epochs based on heart rate variability, respiration rate variability, and binary actigram. Total sleep time (TST), sleep onset latency (SOL, determined as the duration from being present in bed with lights-out to the first five minutes of consecutive sleep), wakefulness after sleep onset (WASO), and sleep efficiency (SE is commonly defined as the ratio of a person’s total sleep time to time spent in bed, which refers to the amount of time a person is actually asleep during the time spent trying to sleep) were obtained of each night. Average values across the nights measured were used for analyses. The possible conditions which might affect the measurement, such as children and pets in the bedroom were recorded in the sleep diary. A research assistant visits each participant’s home to set up the system before measurements started. The sleep data was validated by the piezoelectric system carried out against two-night polysomnography measurement (31 participants with insomnia complaints, aged 51.8 ± 8.4 years, BMI = 30.9 ± 4.8) ^5^.

### 2.4. Exercise intervention

Exercise was prescribed according to the recommendations of the American College of Sports Medicine ^23^. Nordic walking combined with other aerobic exercises was performed 30 to 60 minutes per session, 3 to 5 sessions per week, at the intensity level of 60 – 75% of estimated maximum heart rate. Two experienced exercise trainers were responsible for the instruction and supervision of exercise training once a week and in the other sessions, the participants exercised independently. In the beginning of each week, the trainer informed the participant of the duration and intensity (heart rate interval) of each exercise session ^5^.

### 2.5. Diet intervention

Specific individualized diet program was developed after baseline assessments of each participant’s current dietary intakes (based on three-day food diary) and body weight. The suggested diet contained the following relative macronutrient composition of energy intake: 40% carbohydrate with < 5% sucrose, 40% fat (saturated fatty acid (SAFA) 10%, monounsaturated fatty acid (MUFA) 15 – 20%, polyunsaturated fatty acid (PUFA) 10%) and 20% protein. Overweight/obese participants were advised to moderately reduce their total energy intake (by 300 to 500 kcal per day for the first three months) with guidance on the proportion of macronutrients to be consumed. The target was to reduce body weight by 3 kg in the first three months of the intervention. Detail information was given in previous paper^4^.

### 2.6. Control group

Control group participants were instructed to maintain their habitual, pre-recruitment lifestyle during the intervention. They were invited to a lecture explaining the purpose of the group. After 6-month study period, they were given an opportunity to participate in the exercise plus dietary intervention program for 3 months.

### 2.7. Serum sample collection

Blood samples were collected in the morning between 7:00 and 9:00 a.m. after overnight fasting at baseline and after 6-month intervention from all subjects. Serum was extracted by centrifugation and stored immediately at -80 °C until analyzed.

### 2.8. Metabolomics assessment

#### 2.8.1. Sample preparation

Serum sample (100 μL) was first thawed in room temperature, then mixed with methanol-chloroform (300 uL 3:1, v/v) solvent and L-2-chlorophenylalanine (10 μL, 0.3 mg/ml in water) for metabolites extraction, and then kept at -20°C for 10 min. After centrifugation at 15000 g for 10 min, 300 μL of supernatant was obtained and dried completely under nitrogen. Next, 80 μL methoxyamine (15 mg/mL in pyridine) was added to the vial, vortexed for 30 s and kept at 37°C for 90 min followed by 80 μL BSTFA (1 % TMCS) and 20 μL n-hexane at 70°Cfor 60 min.

Quality control (QC) samples for serum were prepared by pooling equal aliquot of each sample and then were pretreated as serum samples, respectively.

#### 2.8.2. Gas Chromatography Time-Of-Filght Mass Spectrometry (GC-TOF-MS) analysis

Each 1 μL aliquot of the derivatized solution was injected in splitless mode into the Pegasus 4D GC–TOF-MS system (LECO Corporation, St Joseph, MI, USA). Separation was carried out on a non-polar DB-5 capillary column (30 m × 250 μm I.D., J&W Scientific, Folsom, CA), with high purity helium as the carrier gas at a constant flow rate of 1.0 mL/min. The GC temperature programming was set to begin at 90°Cand hold for 0.2 min, and then followed by 10°C/min oven temperature ramps to 180°C, 30°C/min to 240°C, 20°C/min to 280°C, and a final 11 min maintenance at 280°C. The temperature of injection and ion source was set to 280°Cand 220°C, respectively. Electron impact ionization (-70 eV) at full scan mode (m/z 30-600) was used, Solvent delay: 7.6 minutes,with an acquisition rate of 100 spectrum/second in the MS setting.

### 2.9. Data processing and statistics

IBM SPSS statistics version 24 (SPSS, Inc., Chicago, IL, USA) was used to perform statistical analysis. All data were checked for normality by Shapiro–Wilk’s W-test and for homogeneity by Levene’s test before each analysis. Natural logarithm transformations were performed on non-normally distributed data. Spearman correlation analysis was performed with change percentage of metabolites and change percentage of parameters related to sleep. Between-group differences were evaluated by repeated measures ANOVA, followed with Sidak corrections for multiple comparisons. All tests were two sided with significance level p < 0.05.

To account for heteroscedasticity data were normalized by log2-transformation. Multivariate statistical analysis and univariate analysis were used for metabolomics data analysis.

The acquired MS data from GC− MS were analyzed by ChromaTOF software (v 4.34, LECO, St Joseph, MI). The data sets resulting from GC-MS were separately imported into SIMCA-P+ 14.0 software package (Umetrics, Umeå, Sweden). Principle component analysis (PCA) and (orthogonal) partial least-squares-discriminant analysis (O) PLS-DA were carried out with SIMCA-P and differential metabolites were based on variable importance in the projection (VIP) > 1.0 and student *t* test p < 0.05. In the present study, a 7-fold cross validation procedure and 200 random permutations tests were performed to avoid overfitting of the supervised OPLS-DA models. The intensity of each metabolite was used as final results.

## 3. Results

Characteristics of the study participants are shown in **Table 1**. Briefly, during the 6-month intervention period, the control group had increased their body weight and fat mass (FM) significantly, while both intervention groups had no significant changes in body weight, FM, and total energy intake.

**Table 1.** Characteristics of subjects (mean ± SD)

|  | Exercise (n = 24) |  | Diet (n = 27) |  | Control (n = 21) |  | Between-group |  |
| --- | --- | --- | --- | --- | --- | --- | --- | --- |
|  | Baseline | 6 months | Baseline | 6 months | Baseline | 6 months | 1 vs 3 | 2 vs 3 |
| Age (years) | 51.2(9.7) | -- | 51.7(9.3) | -- | 50.7(9.3) | -- |  |  |
| Weight (kg) | 92.3(14.6) | 92.5(13.9) | 93.8(11.8) | 92.7(11.9) | 93.1(17.2)<sup>c</sup> | 94.4(17.8) | 0.176 | 0.005 |
| BMI | 29.3(4.0) | 29.4(3.8) | 29.4(3.7) | 29.0(3.7) | 29.2(4.4) | 29.4(4.6) | 0.612 | 0.047 |
| Total fat mass (kg) | 27.3(9.5) | 27.2(9.0) | 27.4(8.4) | 26.8(8.6) | 28.0(9.7)<sup>c</sup> | 28.9(10.7) | 0.180 | 0.027 |
| Triglycerides (mmol/L) | 1.7(0.7) | 1.8(0.9) | 1.6(0.6) | 1.5(0.6) | 1.5(0.7) | 1.4(0.6) | 0.104 | 0.867 |
| Energy intake (kcal) | 2285.2(561.6) | 2043.1(643.6) | 1981.2(372.1) | 1877.1(487.9) | 2223.5(410.1)<sup>c</sup> | 1920.9(463.7) | 0.588 | 0.606 |
| VO2max | 29.3(9.3)<sup>a</sup> | 31.4(9.3) | 27.2(9.6)<sup>b</sup> | 29.6(10.8) | 30.9(7.9) | 30.4(8.0) | 0.086 | 0.053 |
| Total energy expenditure (MET min/day) | 2349.9(224.8) | 2335.5(235.3) | 2346.1(215.9) | 2398.8(234.9) | 2341.1(242.8) | 2322.6(233.7) | 0.887 | 0.282 |

a: exercise 0m compare with exercise 6m; b: diet 0m compare with diet 6m; c: control 0m compare with control 6m, $P < 0.05$ .
Between-group (1: exercise, 2: diet, 3: control) differences were evaluated by repeated measures ANOVA, followed with Sidak corrections for multiple comparisons.

Of the sleep quality related variables, SOL decreased (p < 0.05) after exercise and SE increased (p < 0.05) after diet intervention. No significant changes in sleep-related parameters were found in the control group (**Fig. 1**).

**Figure 1.**
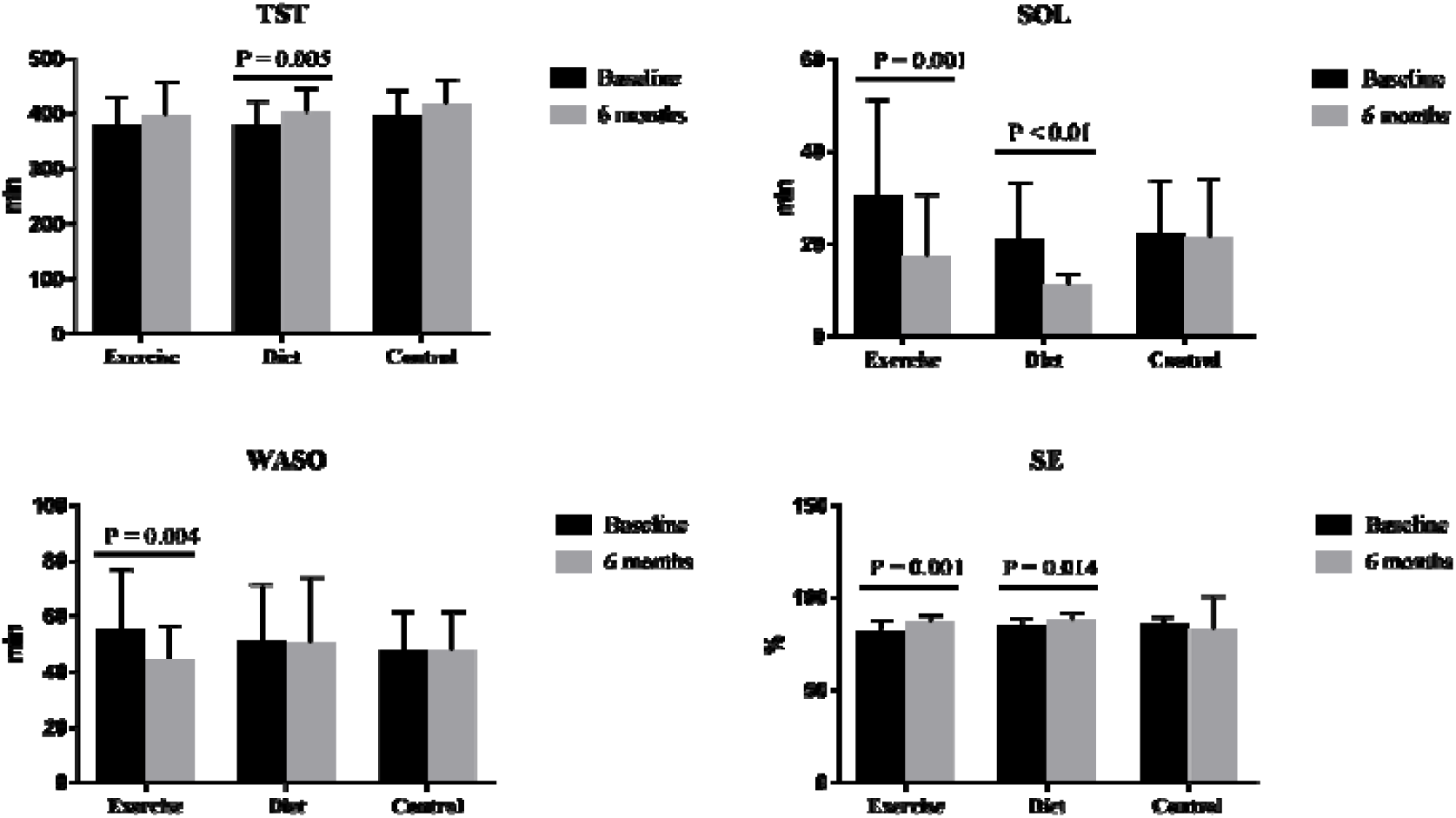
The change of sleep parameters after intervention. TST means total sleep time, SOL means sleep onset latency, WASO means wakefulness after sleep onset, SE means sleep efficient.

A total of 210 known metabolites in serum among different groups were observed. The results show a clear separation between the baseline and follow-up (exercise (R2Y = 0.959, Q2 = 0.675), diet (R2Y = 0.969, Q2 = 0.823) and control (R2Y = 0.981, Q2 = 0.828)) (Supplementary **Fig. S1**). From **Fig. S1B, 2D** and **2F**, the calculated R2 and Q2 values in permutation tests are lower than the original ones and the Q2 intercept on the vertical axis was less than zero. Therefore, the model is considered valid.

The significant metabolites in serum were screened according to the VIP value larger than 1 and p value (p < 0.05) from the *t*/test adjusted by false discovery rate (FDR). There were 21 metabolites affected by exercise intervention, 33 metabolites by diet intervention and 5 in the control group (VIP > 1 and p < 0.05). All metabolites showed greater than 2-fold change between baseline and 6-months, except acetanilide in the exercise and control groups (**Table S1**). The significantly changed metabolites in all groups are shown in **Fig. 2A, 2B and 2C. Figure 2D** shows 2 unique metabolites altered in exercise group, 14 in diet group and 0 in control group.

**Figure 2.**
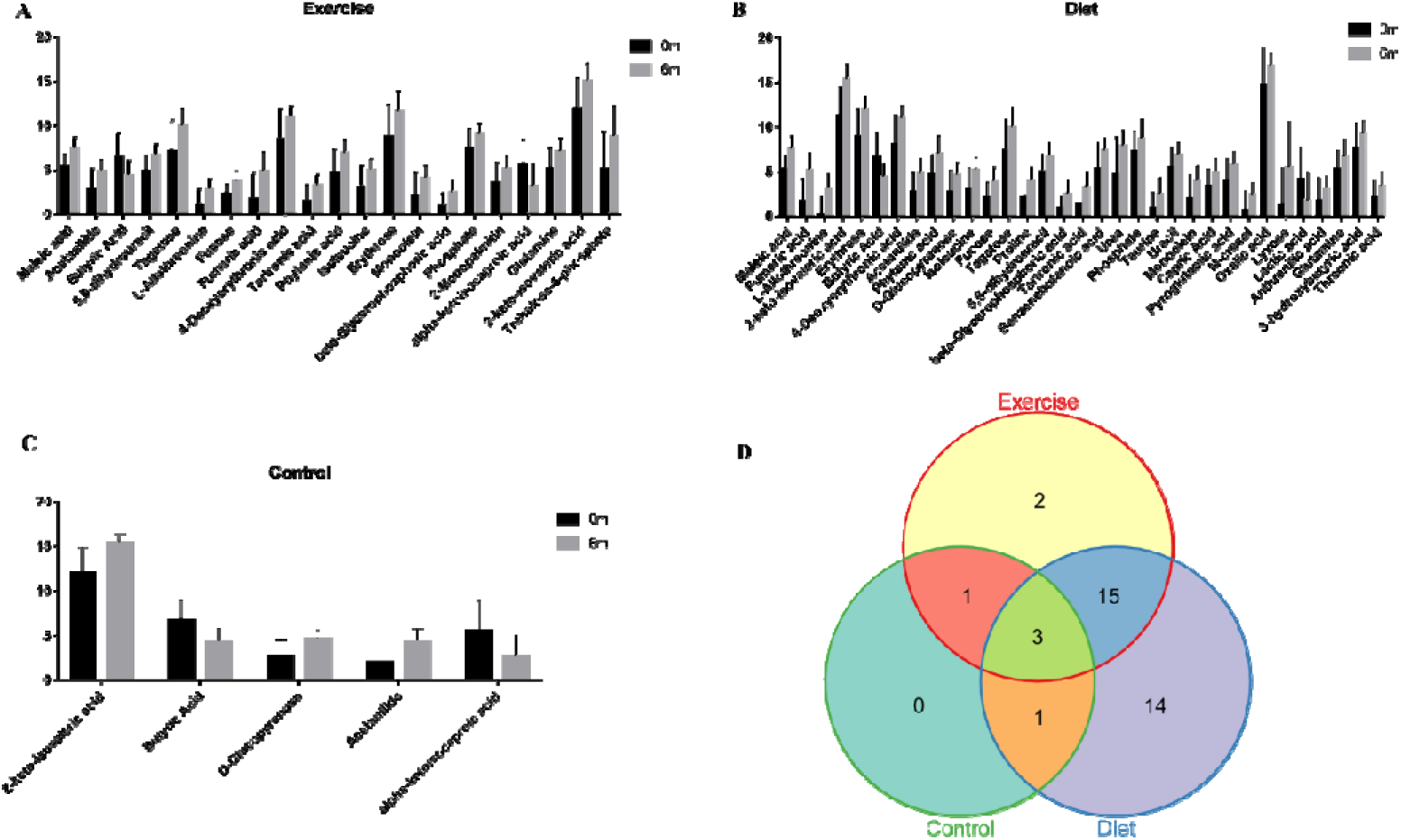
The significant altered metabolites after intervention and venn figure of these metabolites in three groups. A means significant altered metabolites in exercise group, B means significant altered metabolites in diet group, C means significant altered metabolites in control group, and D showed common metabolites and separate metablites in three groups. Y-axis means standard data after log2 tranformation.

We next assessed whether these metabolites were associated with changes in sleep parameters in each individual (**Fig. 3**). We found that decreased alpha-ketoisocaproic acid was correlated with improved SE in the exercise group (r = -0.52, p = 0.026). In addition, despite of no significant association between phosphate and SE, 67% of the participants showed increased phosphate levels after exercise intervention. In the diet group, change of 3-hydroxybutric acid (r = -0.47, p = 0.025) and D-glucopyranose (r = -0.54, p = 0.006) correlated negatively with SE. On the other hand, oxalic acid (r = 0.49, p = 0.021), D-glucopyranose (r = 0.43, p = 0.048), 4-deoxyerythronic acid (r = 0.60, p = 0.004) and tagatose (r = 0.51, p = 0.016) correlated positively with change of SOL, and 2-keto-isovaleric acid (r = 0.45, p = 0.029) correlated with TST in the diet group.

**Figure 3.**
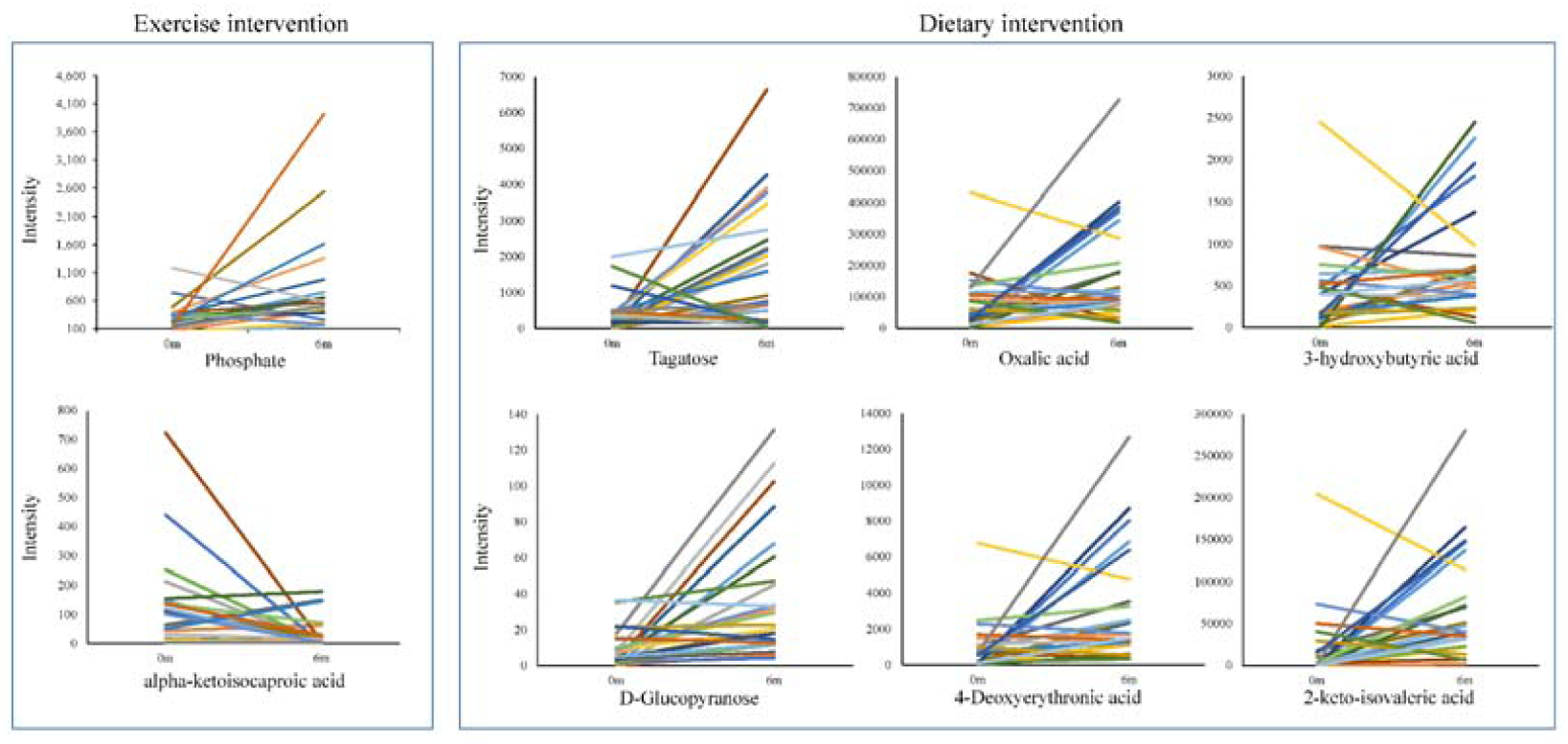
The intensity of significant changed metabolites in individuals between baseline and 6-month follow-up in exercise and diet groups. Each line represents one individual.

## 4. Discussion

In this study, the effects of exercise and dietary intervention on serum metabolome were assessed in men with insomnia symptoms. Twenty-one metabolites changed significantly after exercise intervention and thirty-three metabolites after dietary intervention. Five metabolites changed significantly in the control group during the same period.

Sleep efficiency (SE) is commonly defined as the ratio of a person’s total sleep time to time spent in bed ^24^. Early study have shown that sleep timing is associated with a large number of metabolites across a variety of biochemical pathways ^25^. Widespread changes in circulating metabolites, including reduction of carbohydrates and increased levels of certain lipids and amino acids have been associated with sleep restriction and sleep deprivation ^16, 17, 26^. However, such results may not reflect the alteration of circulating metabolites after exercise and dietary intervention in patients with insomnia symptoms. In this study, we found that concentrations of 21 metabolites changed significantly in subjects with insomnia symptom after exercise intervention. These metabolites were mainly related to glycerolipids metabolism and carbohydrates metabolism. We found that the decreased alpha-ketoisocaproic acid (KIC) was correlated with improved SE after exercise intervention. To our knowledge no previous study has linked exercise and KIC with sleep quality. KIC is metabolic intermediate in the metabolic pathway for L-leucine ^27^, which via leucine dehydrogenase and/or branched chain amino acid transferase at the expense of NH3, in a variety of tissues including skeletal muscle ^28^. An early study by Funchal et al. ^29^ showed that KIC increases phosphorylation of intermediate filament proteins from rat cerebral cortex by mechanisms involving Ca2+ and cAMP. Another study found that KIC and leucine provoke mitochondrial bioenergetic dysfunction in rat brain ^30^. Thus, we speculate KIC and changes in the amino acid metabolism may play a role in exercise induced improvement in SE.

Phosphate was another metabolite associated with the change of sleep quality after 6-month exercise intervention. Phosphate is an essential mineral component of the human body involved on bone and cell membranes, molecules adenosine triphosphate, nicotinamide adenine dinucleotide, cyclic adenosine monophosphate and cyclic guanosine monophosphate metabolism ^31^. Therefore, its dysregulation can affect the functionality of almost every organ system ^32^. Phosphate balance is maintained by the kidneys and to some extent by the gastrointestinal tract in healthy adults, regulated by vitamin D, parathyroid hormone and phosphatonins such as fibroblast growth factor-23 (FGF23) ^31, 33^. Intrinsic factors including circadian rhythm, age and sex, and genetics, including single nucleotide polymorphisms (SNPs) are correlated with serum phosphate and mutations in the sodium–phosphate cotransporters and other hormone including dopamine, Angiotensin II and insulin like growth factor ^34^. Patients with chronic kidney diseases showed that a higher sleep quality score was associated with higher phosphate levels ^35^. Phosphate increased in our study after 6-month exercise intervention in patients with insomnia, consistent with previous study, which suggest appropriate concentration of phosphate is important for sleep ^35^.

The effect of dietary intervention were more pronounced on sleep onset latency (SOL), which is the duration from being present in bed with lights-out to the first five minutes of consecutive sleep. We found that 33 metabolites were changed after dietary intervention. Particularly, increased oxalic acid, D-glucopyranose, 4-deoxyerythronic acid and tagatose are associated with improved SOL after dietary intervention. These metabolites are mainly related to purine metabolism, glucose metabolism, galactose metabolism.

Oxalic acid (oxalate) is derived from diet, degradation of ascorbate, and produced by the liver and erythrocytes. Oxalic acid has been shown to be reduced in both rat and human following sleep restriction, and recover to near baseline levels after sleep restriction ^36^. In the present study, oxalic acid increased after 6-month diet intervention, supporting the notion that oxalic acid is a potential biomarker for sleep quality. In addition, 3-Hydroxybutyric acid (beta-hydroxybutyrate, 3-HB) which is an indicator of the status of hepatic fatty acid oxidation ^37, 38^, has been shown to increase in subcutaneous adipose after sleep loss in humans ^39^. Serum 3-HB was also increased after diet intervention in patients with insomnia symptoms and was proposed to be related to the enhanced transport of plasma free fatty acid into the liver mitochondria ^40^. We also found increased D-glucopyranose after 6-month diet intervention. D-glucopyranose (glucose) is a monosaccharide containing six carbon atoms and an aldehyde group and is therefore referred to as an aldohexose which is a primary source of energy for living organisms. Abnormal glucose metabolism have been linked to disturbances of different aspects of sleep, including sleep duration, quality, respiratory function during sleep, and circadian timing ^41^. Both short and long sleep durations are associated with worse glucose control in people with diabetes ^42^. Recently, *Kay.et al*., showed that good sleepers, but not insomnia patients, had lower whole-brain glucose metabolism during recovery NREM sleep compared to baseline and individuals with insomnia. ^43^.

In addition to oxalic acid, 3-HB and D-glucopyranose, we found increased tagatose and 2-keto-isovaleric acid after 6-month diet intervention. Tagatose (D-tagatose) has been associated with improved glucose control, weight loss and elevated high-density lipoprotein cholesterol ^44^. In addition, 2-keto-isovaleric acid (KIV) arises from the incomplete breakdown of valine, which is one of branched-chain amino acids. KIV accumulate in Maple syrup urine disease (MSUD) patients, which is a metabolic disorder caused by a deficiency of the branched-chain alpha-keto acid dehydrogenase complex (BCKDC) ^45^. However, no study has investigated the effect of tagatose and KIV on insomnia.

Our study has both strengths and limitations. The study participants were assigned to intervention and control groups at random, and sleep was measured using non-contact sleep monitoring system. Exercise training program was prescribed according to the recommendations of the American College of Sports Medicine and was supervised by experienced trainers. Diet intervention was individualized according the subjects body weight and caloric intake, and the duration of exercise and diet interventions were relatively long. The limitations include a small number of subjects and a relatively high drop-out rate in the control group, which may have affected the study results.

## 5. Conclusion

In conclusion, exercise and diet intervention in patients with insomnia symptoms was associated improved sleep quality and changes in serum metabolites involved in glycerolipids metabolism, carbohydrates metabolism, purine metabolism, glucose metabolism. These results suggest circulating metabolites potentially involved in the exercise and diet induced improvement of sleep quality in patients with insomnia symptoms.

## Data Availability

Availability of the data will follow the data management principles for research at the University of Jyväskylä https://www.jyu.fi/tutkimus/tutkimusaineistot/rdmenpdf. Established researchers wishing to collaborate will be given access to the de-identified data following
approval of a signed research proposal.

## Acknowledgments

This study was supported financially by the by the Finnish Funding Agency for Technology and Innovation (TEKES2206/31/2010), the 111 Project (B17029) of Shanghai Jiao Tong University, the Shanghai Jiao Tong University Zhiyuan Foundation (Grant CP2014013), and China Postdoc Scholarship Council (201806230001).

## Authors’ contributions

The authors’ contributions are as follows: SC is the principal investigator (PI). She designed the study and oversaw the implementation of the project, trained the researchers, supervised the doctoral students, and participated in the data collection, analyses and interpretation, and editing the manuscript. XZ, XW and PW drafted the manuscript. SL, XO and XT participated in the data analyses, interpretation and commenting the manuscript.

